# Establishment of Local Diagnostic Reference Levels for Trunk Computed Tomography Examinations at Governmental Hospitals in the Gaza Strip: A Cross-Sectional Study

**DOI:** 10.64898/2025.12.30.25343210

**Authors:** Amjad Ayyad, Yasser Alajerami, Ahmed Najim, Husam H. Mansour, Muhammad Khalis Abdul Karim, Fahad Alghamdi, Kinan Mokbel

## Abstract

**Objective:** To establish the local diagnostic reference levels (LDRLs) for trunk multi-slice CT (MSCT) examinations in the Gaza Strip, Palestine.

**Method:** A cross-sectional study of adult oncology patients undergoing trunk MSCT at two governmental hospitals in Gaza Strip; Al Shifa Medical Complex (SMC) and Al Aqsa Hospital (AMH), using an adapted dose survey booklet. Data collected from July 2019 to March 2020 included patient characteristics, volumetric CT dose index (CTDI_vol_) and dose length product (DLP). Descriptive, univariate and multivariate analyses identified key factors affecting radiation dose, and the coefficient of variation between scanner and software-derived dose values was also determined.

**Results:** A total of 170 trunk CT examinations were analysed (57.1% SMC, 42.9% AMH). 73% were female; the mean age of the participants is 53.1±15.8 years, and the mean body mass index (BMI) was 30±6.1. The estimated LDRLs for trunk CT were 13 mGy for CTDI_vol_ and 1010.4 mGy·cm for DLP. There was notable variation between hospitals in CTDI_vol_ and DLP (*p*<0.001). At SMC, factors such as tube current, peak kilovoltage, scan length, pitch and BMI significantly affected dose indices. In contrast, at AMH, the main influences were tube current and scan length. CTDI_vol_ had a greater impact on DLP than scan length at both locations.

**Conclusion:** LDRLs for trunk CT scans in the Gaza Strip were established and found to be generally comparable to international benchmarks. Notable variation in doses between hospitals indicates potential for improvement through standardising protocols, managing scan lengths and using techniques tailored to patient size.

## Introduction

Computed tomography (CT) plays a crucial role in modern diagnostics and follow-up care. The rapid advances in Multi-Slice Computed Tomography (MSCT) scanners make it a valuable diagnostic tool, leading to a surge in CT exam requests [1]. However, it accounts for a significant portion of medical radiation exposure relative to other imaging methods, highlighting the importance of optimisation as a key aspect of radiological governance [2-4]. Technological improvements in CT have reduced radiation dose, but significant hospital-level differences remain [5-7]. To address this and improve dose optimisation, the International Commission on Radiation Protection (ICRP) introduced Diagnostic Reference Levels (DRLs) [8].

DRLs are widely recognised as a practical optimisation tool for identifying unusually patient dose levels for a given examination and for prompting protocol review and standardisation [8]. In a dose survey, the third quartile value of each examination, where 75% of the data is below it, is used as the DRL acceptance criterion [9, 10]. Regional and international DRLs are inadequate because of differences in region-specific training, equipment and populations used to establish them [11]. Many countries have established local and national DRLs for radiological examinations, with reviews showing substantial dose reductions (16%-30%) from their use [12-14].

Patient dose can be minimised through proper protocol selection, scanning parameters and patient positioning. Developing countries like Palestine face limited equipment, poor maintenance and weak radiation protection. Radiological centres in Gaza may face challenges related to the consistency of equipment calibration, optimisation of radiation exposure and standardisation of imaging protocols. Data on CT exam frequency is usually missing, preventing identification of hospitals with abnormal doses. Establishing a local diagnostic reference level (LDRL) is essential for dose optimisation. The LDRL can guide MSCT procedures to manage and optimise dose, ensuring it is appropriate for clinical needs [15]. Many organisations support the use of DRLs as a dose-optimisation tool [16].

This study aimed to establish the LDRL for trunk multi-slice CT (MSCT) examinations in the Gaza Strip. Secondary objectives included quantifying key dose indicators, comparing mean dose values with regional and international benchmarks, assessing inter-hospital variation in dose metrics, identifying the principal scan-parameter predictors of volumetric CT dose index (CTDI_vol_) and dose-length product (DLP) to support dose optimisation, and evaluating the agreement and variability between scanner console-reported dose values and software-generated dose estimates.

## Materials and Methods

### Study Design and Ethics Approval

In a cross-sectional study, data were obtained from 458 patients at two hospitals in the Gaza Strip, Al Shifa Medical Complex (SMC) and Al Aqsa Martyrs Hospital (AMH). Ethical approval was granted by the Helsinki Committee (HC) for Ethical Approval, Palestinian Health Research Council (PHRC), Gaza Strip, Palestine (approval number PHRC/HC/572/19; 17 June 2019). Administrative approval was obtained from the Ministry of Health, State of Palestine (Human Resources Development Directorate; correspondence number 381554; 17 Oct 2019). Informed consent was obtained from all participants before participation. The specifications of the CT machines are shown in Table 1. Data was collected between July 2019 to March 2020.

**Table 1:** Specifications of machines in each hospital. SMC: Al Shifa Medical Complex; AMH: Al Aqsa Martyrs Hospital.

| Hospital | Manufacturer | Model | Slices | Manufacturing Year | Installation Year |
| --- | --- | --- | --- | --- | --- |
| SMC | Philips Health Care | Ingenuity Core<sup>128</sup>™ | 128 | Oct-2013 | May-2015 |
| AMH | General Electric Health Care | Optima CT540 | 16 | Feb-2016 | March-2017 |

### Inclusion and Exclusion Criteria

The study inclusion criteria were adult oncology patients aged ≥19 years who were registered and underwent MSCT of the trunk at AMH or EGH. Exclusion criteria were non-oncology status, MSCT protocols other than the trunk examination, MSCT performed outside AMH/EGH and attendance in a wheelchair or on a trolley (excluded on ethical grounds).

### Data Collection Tools

Data was collected for each trunk CT examination performed for the specified clinical indication (oncology follow-up or lymphoma staging) to standardise data capture and reduce protocol-related heterogeneity, as different clinical indications may require different acquisition protocols, even when imaging the same anatomical region. Dose and scan-parameter data were recorded at two points: first, during protocol selection, and second, immediately after the examination. This enabled cross-checking against the scanner’s dose report at the end of each scan.

Patient demographic information, acquisition parameters and dose indices were collected using an adapted, self-administered Canadian CT dose survey booklet (Supplement, S1). The following parameters were extracted from the CT console: tube potential (kV), tube current (mA), rotation time (s), tube current–time product (mAs), beam collimation (N×hcol), table feed, slice thickness, scan length, pitch, CTDI_vol_ and DLP. The effective dose values were calculated across the surveyed hospitals primarily because of differences in the DLP values used for the examinations studied, and for general comparison with other imaging procedures, but was not a primary outcome, as the study focused on establishing LDRLs for dose optimisation [17].

CT-Expo dose calculation software (version 2.3.1; Germany) was subsequently used to validate and compare the dose values reported by the CT scanners, thereby assessing the reliability of console-displayed dose metrics. When scanner- and protocol-specific inputs are provided, CT-Expo outputs CTDI_vol_ and the corresponding DLP and estimates effective dose (and organ doses) using ICRP publications 60 or 103 tissue-weighting factors [18, 19].

### Statistical Analysis

Microsoft Excel (Microsoft, Redmond, WA, USA) and SPSS Statistics version 23.0 (SPSS Inc., Chicago, IL, USA) were used for data management and statistical analysis. The initial sample comprised 210 examinations, of which 10% were excluded as part of a pilot phase. To minimise the influence of outliers, the upper and lower 5% of outcome values were excluded using pairwise trimming, reducing the analytical dataset from 190 to 170 examinations. This 10% trimming strategy aligns with ICRP guidance [15].

Descriptive statistics (frequency tables, scatterplots and figures) were used to summarise the data. Effective dose was estimated from the DLP using ED (mSv)=K×DLP, where K=0.015 for trunk (chest-abdomen-pelvis) CT [20]. Diagnostic reference levels (DRLs) for CTDI_vol_ and DLP were defined as the 75th percentile (third quartile) of their distributions. Between hospital differences in outcome measures were assessed using an independent samples t-test. Pearson’s correlation coefficient was used to examine associations between scan parameters and outcome variables (CTDI_vol_ and DLP). Multiple linear regression was performed to identify predictors of CTDI_vol_ and DLP. Statistical significance was set at *p*<0.05 with 95% confidence intervals. In addition, the coefficient of variation between scanner-reported and software-calculated dose values was computed.

## Results

### Patient characteristics and chest CT scan parameters

During the second half of the data collection period, the CT scanner at EGH malfunctioned. Consequently, EGH cases were transferred to AH, enabling data collection to continue during regular working hours and on standard working days. Overall, trunk CT examinations were distributed across the surveyed hospitals as follows: 97 cases (57.1%) were performed at SMC and 73 cases (42.9%) at AMH. Participants’ characteristics, as shown in Table 2, indicate that 27% are male, and 73% are female. The average age of the patients examined is 53.1±15.8 years, and the average body mass index (BMI) is 30±6.1.

**Table 2:**
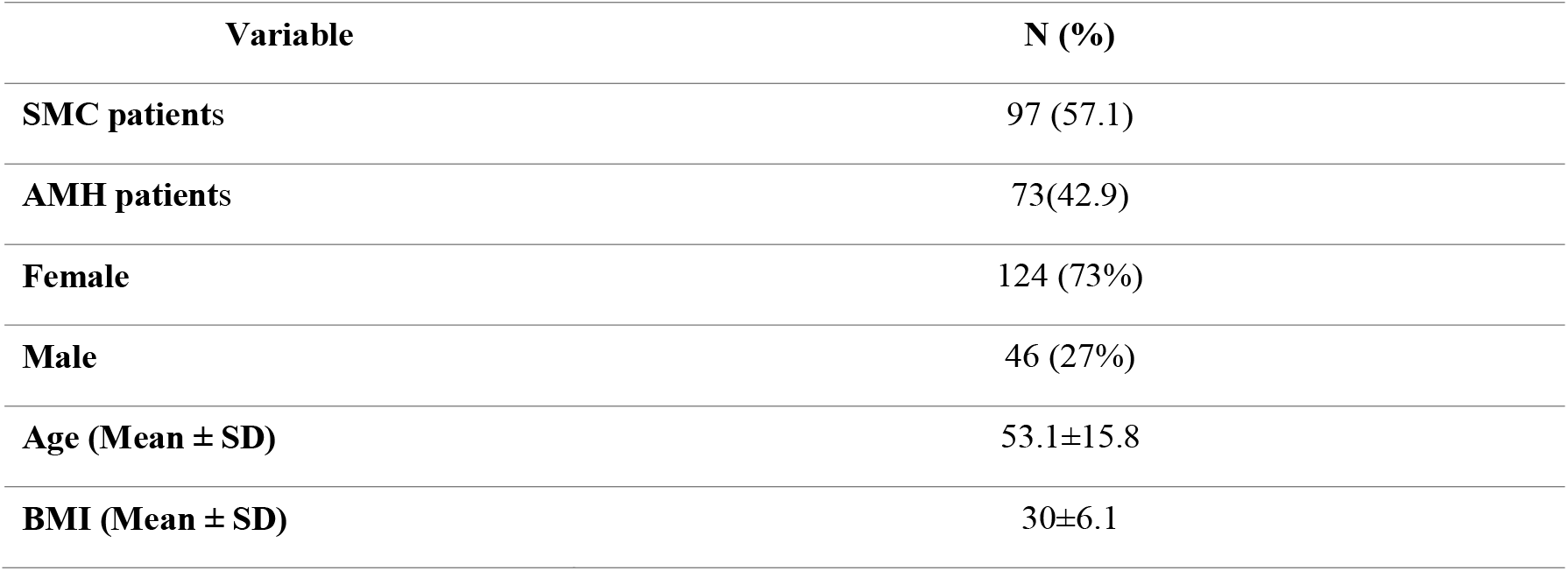
Distribution of Participants Characteristics (n=170). SMC: Al Shifa Medical Complex; AMH: Al Aqsa Martyrs Hospital; BMI: body mass index; SD: standard deviation.

### Radiation parameters

The factors affecting CTDI_vol_ and DLP values at the two hospitals are illustrated in Table 3. The mean peak kilovoltage in SMC is 120kVp (range: 100-140kVp), whereas in AMH it is fixed at 120kVp. Regarding tube current, the mean in SMC is 285.3mA (range: 266-400mA), whereas in AMH it is 515.9mA (range: 500-733.3mA). Additionally, the mean tube current-time product is 214 mAs (range: 200-300mAs), whereas in AMH it is 309.59mAs (range: 300-400mAs). The mean pitch is 0.99 (range: 0.04-0.83) in SMC, whereas in AMH it is fixed at 1.37. For scan length, the mean is 667mm (range: 570-850mm) in SMC and 708mm (range: 565-951mm) in AMH. The fixed values at the two hospitals do not affect CTDI_vol_ or DLP. The rotation time, number of rows, nominal slice thickness, beam collimation, reconstructed slice thickness and no titling were recorded in all exams. Finally, the mean effective dose values are SMC 15.6 mSv and AMH 12mSv.

**Table 3:** Parameters Affecting the CTDI_vol_ and DLP Values (n=170). SMC: Al Shifa Medical Complex; AMH: Al Aqsa Martyrs Hospital; SD: standard deviation; ‡ (detector Configuration product).

| Variable | SMC | AMH |
| --- | --- | --- |
| | Mean ± SD | Mean ± SD |
| Peak kilovoltage | 120.41±4.98 | 120 (fixed) |
| Tube current (mA) | 285.35±30.95 | 515.98± 59.34 |
| Current time product (mAs) | 214.02±23.21 | 309.59± 35.61 |
| Pitch | 0.99±0.04 | 1.37 (fixed) |
| Scan length (mm) | 667±56.9 | 708±70.9 |
| Rotation time (s) | 0.75 (fixed) | 0.60 (fixed) |
| Number of rows | 64 (fixed) | 16 (fixed) |
| Nominal slice thickness (mm) | 0.625 (fixed) | 1.250 (fixed) |
| Beam Collimation (mm)‡ | 40 (fixed) | 20 (fixed) |
| Reconstructed slice thickness (mm) | 2 (fixed) | 2.5 (fixed) |
| **Gantry Tilt** | 0 (fixed) | 0 (fixed) |
| **Effective dose (mSv)** | 15.6 | 12 |

### Inter-Hospital Dose Variation and Local Diagnostic Reference Levels

The calculated DRL obtained from the two hospitals based on CTDI_vol_ and DLP estimated LDRL for Trunk CT examinations was expressed as the third quartile (75th percentile) of the CTDI_vol_ and DLP values, as shown in Table 4. There were statistically significant differences in CTDI_vol_ and DLP between the two hospitals surveyed. The mean CTDI_vol_ was higher in SMC than in AMH (14.2 vs 11.7 mGy, *p*<0.001). Additionally, the mean DLP was higher in SMC than in AMH (1041.6 vs 799.3 mGy.cm, *p*<0.001).

**Table 4:** Differences in CTDI_vol_ and DLP values across the surveyed hospitals and the estimated local DRL for Trunk CT examinations. mGy: milligrays; mGy.cm: milligrays per centimetre; SMC: Al Shifa Medical Complex; AMH: Al Aqsa Martyrs Hospital; SD: standard deviation; * Statistically significant p<0.05.

| CT Parameters | Hospital | N | Mean±SD | p | 75<sup>th</sup> percentile |
| --- | --- | --- | --- | --- | --- |
| **$CTDI_{vol}$ (mGy)** | SMC | 97 | 14.2±2.5 | <0.001* | 13.0 |
| | AMH | 73 | 11.7±0.5 | | |
| **DLP (mGy.cm)** | SMC | 97 | 1041.6±201.8 | <0.001* | 1010.4 |
| | AMH | 73 | 799.3±68.2 | | |

The 75^th^ percentile values obtained represent CT practice in the Gaza Strip and can be used to compare with related regional surveys, recommended standards and other countries. The calculated DRLs for the two hospitals are based on CTDI_vol_ of 13 mGy and DLP of 1010.4 mGy.cm. The obtained results were compared with DRLs from regional and international countries, including the UK, Canada, the USA, Libya, Saudi Arabia, Singapore, Australia and Japan [21-28]. Furthermore, these values are lower than those in most other countries. The current CTDI_vol_ DRL value is in line with the UK value (13mGy) and lower than the USA values (15mGy). The DLP DRL value (1010.4mGy.cm) is higher than that obtained in the UK (1003 mGy.cm) (Shrimpton et al., 2014) and USA (947mGy.cm). It is also notable that the DRL values for DLP and CTDI_vol_ are higher than the reported values in Singapore (13mGy and 1010.4mGy.cm vs. 12mGy and 823mGy.cm), as illustrated in Figures 1 and 2.

**Figure 1.**
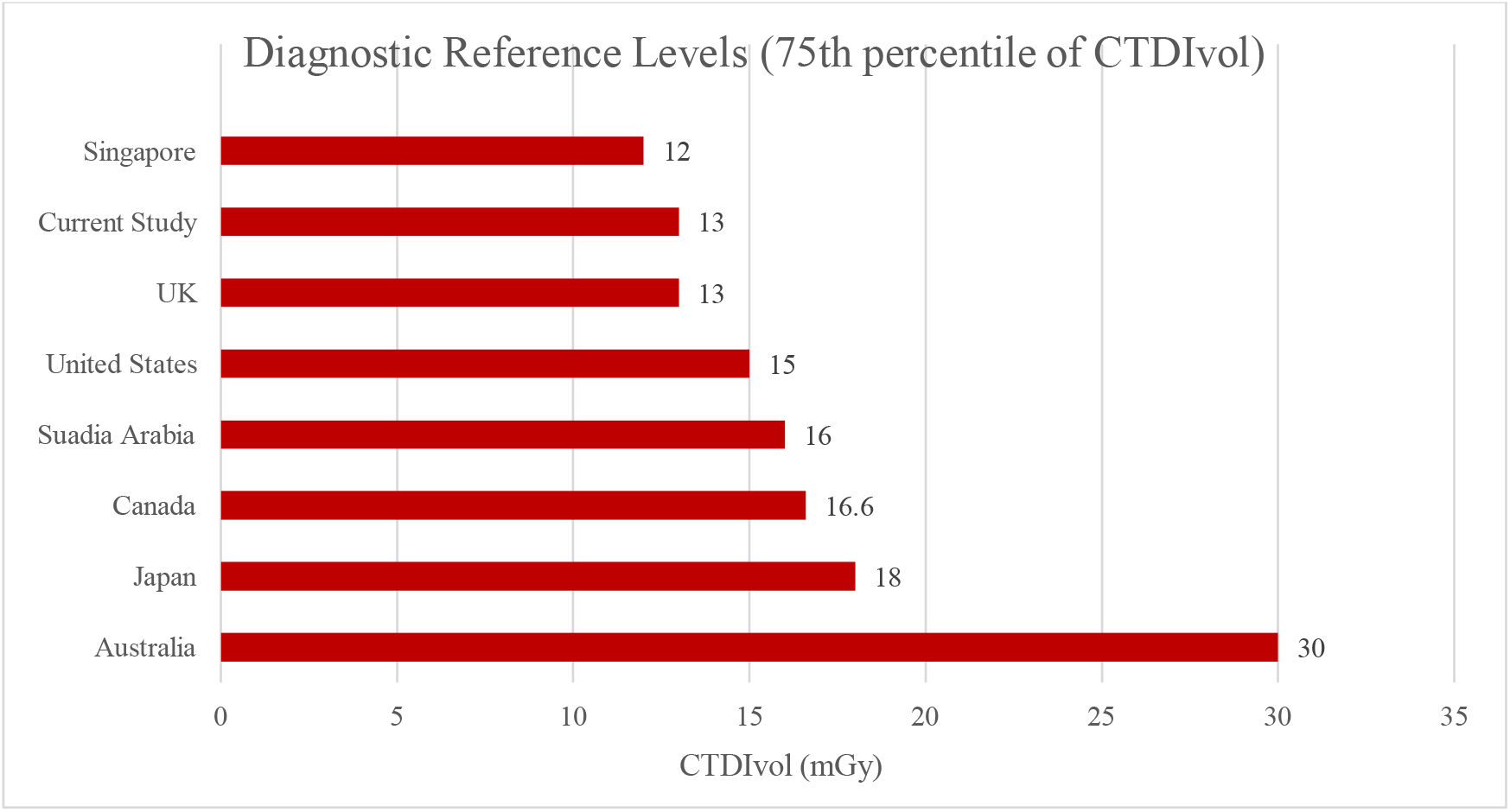
Comparison between local DRL (75^th^ CTDI_vol_) and other countries.

**Figure 2.**
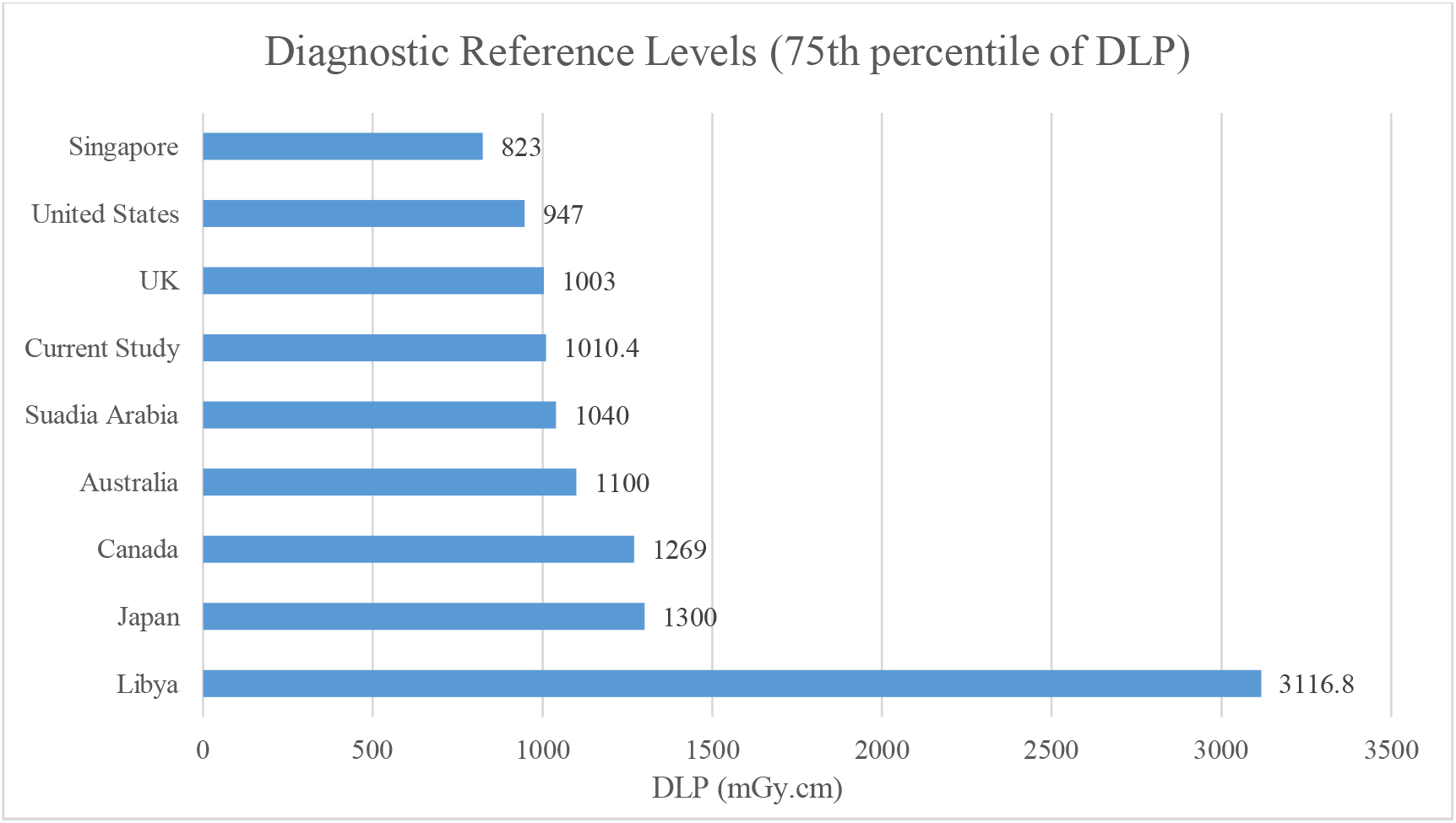
Comparison between local DRL (75^th^ DLP) and other countries.

### Influence of Scan Parameters on Dose Indices

Table 5 presents Pearson correlation analyses between scan parameters and radiation dose indicators across the studied hospitals. At SMC, peak kilovoltage showed a strong positive correlation with CTDI_vol_ (*r*=0.772, *p*<0.001) and a moderate positive correlation with DLP (r=0.623, *p*<0.001), indicating that increases in tube voltage were associated with higher radiation dose metrics. Tube current was also strongly correlated with both CTDI_vol_ (*r*=0.701, *p*<0.001) and DLP (*r*=0.706, *p*<0.001), confirming its substantial influence on patient radiation dose. Pitch showed a statistically significant inverse association with CTDI_vol_ (*r*=−0.358, *p*<0.001) and DLP (*r*=−0.273, *p*=0.007), suggesting that higher pitch values were associated with reduced dose indices. BMI showed moderate positive correlations with CTDI_vol_ and DLP at SMC (*r*=0.469 and 0.375, respectively; *p*<0.001). At AMH, tube current exhibited a strong correlation with CTDI_vol_ (*r*=0.798, *p*<0.001) but weakly with DLP (*r*=0.455, *p*<0.001). At the same time, BMI showed no significant association with either dose metric. Peak kilovoltage and pitch were not analysed at AMH due to fixed protocol settings. Partial correlation analysis showed that DLP was strongly associated with CTDI_vol_ and scan length at SMC (r=0.978 and 0.911; *p*<0.001) and moderately associated at AMH (r=0.650 and 0.636; *p*<0.001). In both hospitals, the association between DLP and CTDI_vol_ was stronger than that with scan length, indicating CTDI_vol_ as the dominant contributor to DLP.

**Table 5:**
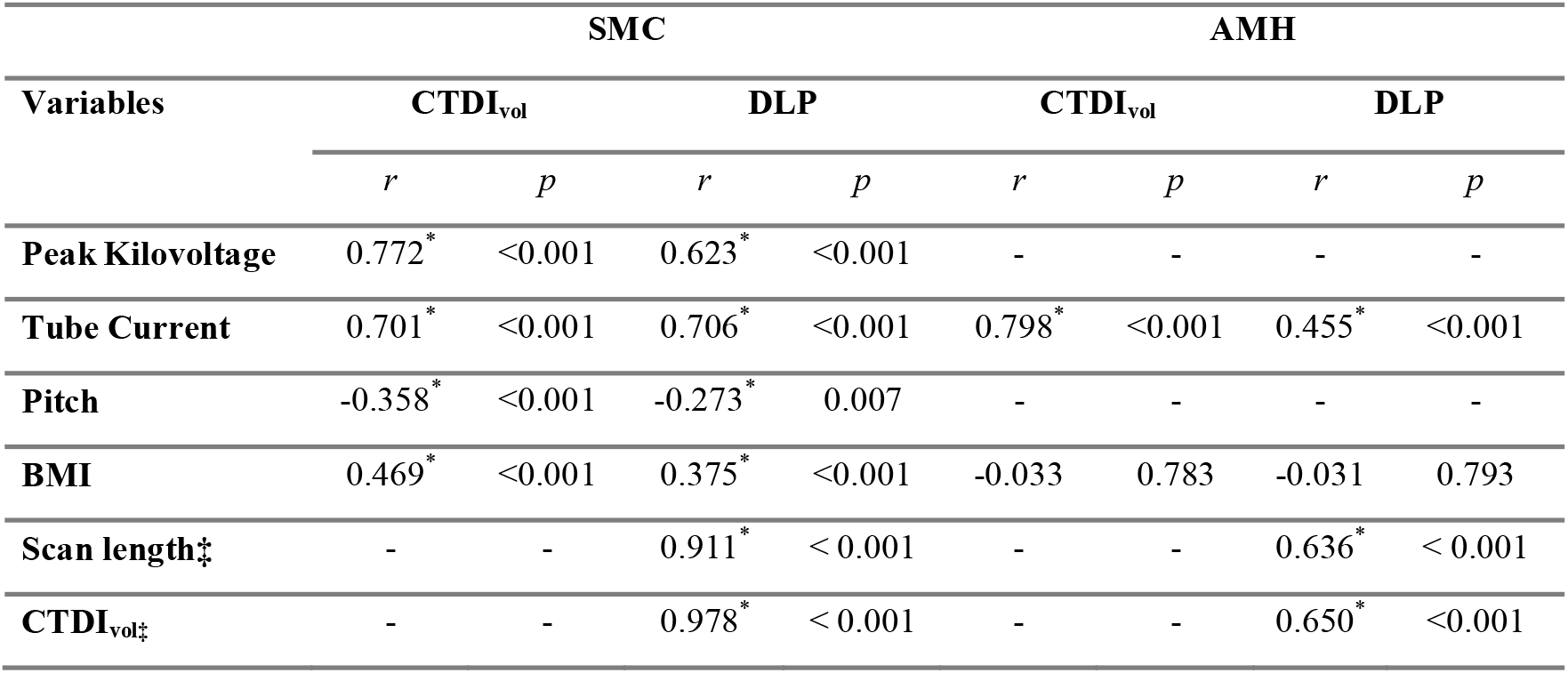
Correlation between the Outcome Variables and the Independent Variables. SMC: Al Shifa Medical Complex; AMH: Al Aqsa Martyrs Hospital; BMI: body mass index; ‡: Partial Correlation between DLP and Each of CTDI_vol_ and Scan Length * Statistically significant p<0.05.

| Variables | SMC |  |  |  | AMH |  |  |  |
| --- | --- | --- | --- | --- | --- | --- | --- | --- |
| | $CTDI_{vol}$ | | DLP | | $CTDI_{vol}$ | | DLP | |
|  | r | p | r | p | r | p | r | p |
| **Peak Kilovoltage** | 0.772* | <0.001 | 0.623* | <0.001 | - | - | - | - |
| **Tube Current** | 0.701* | <0.001 | 0.706* | <0.001 | 0.798* | <0.001 | 0.455* | <0.001 |
| **Pitch** | -0.358* | <0.001 | -0.273* | 0.007 | - | - | - | - |
| **BMI** | 0.469* | <0.001 | 0.375* | <0.001 | -0.033 | 0.783 | -0.031 | 0.793 |
| **Scan length‡** | - | - | 0.911* | < 0.001 | - | - | 0.636* | < 0.001 |
| **$CTDI_{vol}\ddagger$** | - | - | 0.978* | < 0.001 | - | - | 0.650* | <0.001 |

### Beam Collimation, Tube Current Modulation and Protocol Standardisation

The effects of beam collimation on CTDI_vol_ and DLP across the two hospitals are summarised in Table 6. Beam collimation was associated with statistically significant differences in both dose metrics. CTDI_vol_ was higher at SMC with 40 cm collimation (14.2±2.5mGy) than at AMH with 20 cm collimation (11.7±0.5 mGy; *p*<0.001). Similarly, DLP was significantly greater at SMC (1041.6 ± 201.9 mGy.cm) than at AMH (799.3±68.2 mGy.cm; *p*<0.001). For automated tube current modulation (TCM); there were no statistically significant differences in CTDI_vol_ or DLP between TCM used versus not used within either hospital (SMC: CTDI_vol_ *p*=0.640, DLP *p*=0.584; AMH: CTDI_vol_ *p*=0.059, DLP *p*=0.154) (Table 7).

**Table 6:**
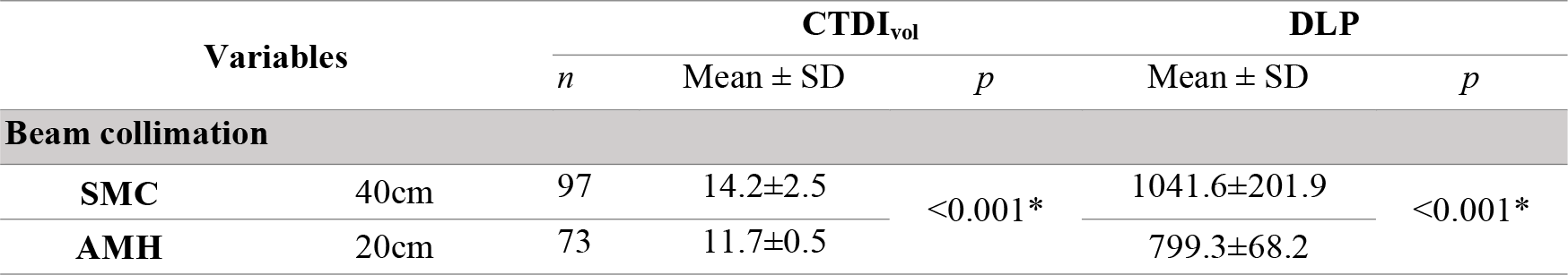
The Effect of Beam Collimation on the Resultant CTDI_vol_ and DLP. SMC: Al Shifa Medical Complex; AMH: Al Aqsa Martyrs Hospital; CTDI_vol_: volumetric CT dose index; DLP: dose length product; n; number of participants from each hospital; SD: standard deviation.

**Table 7:** The Effect of Tube Current Modulation on the Resultant CTDI_vol_ and DLP. SMC: Al Shifa Medical Complex; AMH: Al Aqsa Martyrs Hospital; CTDI_vol_: volumetric CT dose index; DLP: dose length product; n; number of participants from each hospital; SD: standard deviation.

| Variables | | $CTDI_{vol}$ | | | DLP | |
| --- | --- | --- | --- | --- | --- | --- |
| | | n | Mean ± SD | p | Mean ± SD | p |
| **Tube Current Modulation** |  |  |  |  |  |  |
| **SMC** | Used | 1 | 13.04 | 0.640 | 930.8 | 0.584 |
| | Not used | 96 | 14.2±2.5 | | 1042.8±202.6 | |
| **AMH** | Used | 6 | 12.89±1.3 | 0.059 | 882.02±131.0 | 0.154 |
| | Not used | 67 | 11.59 | | 791.87±55.7 | |

Multivariable linear regression was performed to identify scan parameters independently associated with CTDI_vol_ and DLP at SMC and AMH (Table 8). At SMC, tube current (B=0.047, 95% CI:(0.042;0.052); *p*<0.001) and peak kilovoltage (B=0.340, 95% CI: (0.311;0.370); *p*<0.001) were significant independent predictors of CTDI_vol_. At AMH, only tube current remained significantly associated with CTDI_vol_ (B=0.007, 95% CI:(0.006;0.008); *p*<0.001). At SMC, tube current (B=3.452, 95% CI:(2.999;3.905); *p*<0.001), peak kilovoltage (B=23.431, 95% CI: (20.722;26.140); *p*<0.001), and scan length (B=1.435, 95% CI:(1.207;1.664); *p*<0.001) were independently associated with increased DLP. At AMH, DLP was significantly influenced by tube current (B=0.475, 95% CI:(0.225;0.724); *p*<0.001) and scan length (B=0.213, 95% CI:(0.002;0.423); *p*=0.048).

**Table 8:** Multivariate Analysis for Factors Affecting CTDIvol and DLP. SMC: Al Shifa Medical Complex; AMH: Al Aqsa Martyrs Hospital; CTDIvol: volumetric CT dose index; DLP: dose length product; B: regression coefficient; CI: confidence interval. * Statistically significant p<0.05, CI= 95%.

| Variable | SMC |  | AMH |  |
| --- | --- | --- | --- | --- |
|  | B (95% CI) | p | B (95% CI) | p |
| **$CTDI_{vol}$** |  |  |  |  |
| **Constant** | -37.44 (-42.9;-31.89) | < 0.05 | 8.146 (7.158;9.133) | < 0.05 |
| **Body mass index** | -0.005 (-0.035;0.025) | 0.735 | 0.006 (-0.005;0.017) | 0.259 |
| **Tube current** | 0.047 (0.042;0.052) | < 0.001* | 0.007 (0.006;0.008) | < 0.001* |
| **Scan length** | 0.000 (-0.003;0.002) | 0.722 | 0.000 (-0.001;0.001) | 0.660 |
| **Pitch** | -2.410 (-6.040;1.219) | 0.190 | - |  |
| **Peak kilovoltage** | 0.340 (0.311;0.370) | < 0.001* | - |  |
| **DLP** |  |  |  |  |
| **Constant** | -3584.565 (-4096.53;-3072.61) | < 0.05* | 386.284 (189.808;582.759) | < 0.05* |
| **Body mass index** | 0.289 (-2.472;3.049) | 0.836 | 0.587 (-1.534;2.709) | 0.582 |
| **Tube current** | 3.452 (2.999;3.905) | < 0.001* | 0.475 (0.225;0.724) | < 0.001* |
| **Scan length** | 1.435 (1.207;1.664) | < 0.001* | 0.213 (0.002;0.423) | 0.048* |
| **Pitch** | -148.433 (-483.347;186.481) | 0.381 | - |  |
| **Peak kilovoltage** | 23.431 (20.722;26.140) | < 0.001* | - |  |

## Discussion

This study established LDRLs for trunk CT exams at two hospitals in the Gaza Strip, with key factors influencing radiation dose. The findings also offer a snapshot of current CT practice and a basis for dose optimisation and future DRL development. Trunk CT exams made up a significant part of CT activity in the surveyed hospitals, mainly for oncology follow-up and lymphoma staging. SMC conducted more exams than AMH, reflecting Gaza City’s concentrated oncology services and cancer trends across the Gaza Strip [29]. The mean BMI shows a mostly overweight to obese group; This high BMI distribution affects dose optimisation, as patient size influences exposure and radiation dose [30].

The calculated LDRLs for trunk CT exams, CTDI_vol_ (13 mGy); DLP (1010.4 mGy.cm), reflect current practice across the two hospitals and are within international ranges. The CTDI_vol_ DRL matches UK values and is lower than US levels, while the DLP DRL is slightly higher than UK and US benchmarks but less than in other regions [23, 27]. Overall, we observed that trunk CT doses in Gaza are not excessive but could be optimised, especially in scan-length control. On the other hand, significant differences in CTDI_vol_ and DLP appeared between SMC and AMH, with SMC showing higher dose indices despite AMH using higher tube current. Variations likely stem from differences in scanner type, operator protocols and scanning parameters. This underscores the multifactorial nature of CT radiation dose, influenced by scanner characteristics, beam collimation, pitch, scan length and protocols. These findings align with prior studies highlighting hospital variability for the same exams yet emphasising the need for local DRLs over international standards [31, 32].

Our correlation and multivariable analyses confirm that tube current and voltage mainly affect CTDI_vol_, especially at SMC, where protocols vary in kVp. The strong positive associations observed are consistent with prior CT research, demonstrating that automated kVp selection reduces CTDI_vol_ compared with fixed kVp protocols while maintaining diagnostic image quality in oncology CT examinations [33, 34]. In addition, tube current is directly linearly related to radiation dose [35]. At AMH, with fixed kVp and pitch, tube current remains the main predictor, indicating limited protocol customisation.

Our fundings demonstrate that scan length was a key predictor of DLP in both hospitals, highlighting its role in total dose. While CTDI_vol_ had a stronger link, excessive or unclear scan ranges can raise patient exposure without additional benefit. Longer scan lengths in surveyed hospitals likely explain the higher DLP compared to international DRLs, such as Singapore [26]. Furthermore, pitch showed an inverse relationship with dose at SMC; however, this effect disappeared in multivariable modelling.

This aligns with modern multislice CT systems, where automatic tube current adjustment compensates for pitch changes, resulting in minimal dose reduction [36]. BMI was significantly associated with dose at SMC but not at AMH, suggesting patient size was not well integrated into protocols at AMH. This underscores the need for BMI adapted protocols to ensure consistent image quality and prevent unnecessary dose increases.

In the present study, beam collimation showed significant differences in CTDI_vol_ and DLP between hospitals, but these are likely due to other protocol factors. Contrary to expectations, higher doses at SMC are better explained by higher kVp, longer scan length and scanner design, rather than by collimation alone. TCM did not show a significant dose reduction in either hospital, unlike the established literature [37]. This is likely due to limited and inconsistent use of TCM in the cohort, possibly due to poor protocol setup, centering or limited familiarity with TCM. The results suggest that optimising TCM requires proper and consistent clinical application.

This study has, however, some limitations. It was conducted at only two hospitals and included only adult oncology patients, which could limit the applicability of the findings to other clinical settings and populations. Furthermore, variations in CT scanner models and fixed protocol parameters restricted the evaluation of certain dose determinants. Also, the cross-sectional design prevents assessment of long-term dose optimisation and removing extreme values may have excluded some high-dose examinations.

## Conclusions

This study established LDRLs for trunk CT examinations at two governmental hospitals in the Gaza Strip, providing the first evidence-based benchmarks for dose optimisation in this setting. Significant inter-hospital variation in CTDI_vol_ and DLP was observed, driven primarily by modifiable scan parameters rather than patient characteristics. Tube current, tube voltage and scan length were identified as the main determinants of radiation dose, with CTDI_vol_ contributing more strongly to DLP than scan length. Although the derived LDRLs are broadly comparable to international values, the findings indicate opportunities for further optimisation, particularly through improved protocol standardisation and scan-range control. The results support the use of LDRLs as a practical tool for dose monitoring, optimisation and the future development of national diagnostic reference levels in Palestine.

## Ethics declarations

Ethical approval was granted by the Helsinki Committee (HC) for Ethical Approval, Palestinian Health Research Council (PHRC), Gaza Strip, Palestine (approval number PHRC/HC/572/19; 17 June 2019). Administrative approval was obtained from the Ministry of Health, State of Palestine (Human Resources Development Directorate; correspondence number 381554; 17 Oct 2019). Informed consent was obtained from all participants before participation. Declaration that all supporting documentation concerning ethical review and patient informed consent is available upon request.

## Data availability

Data supporting the findings of this study are included in the article and Supplementary Materials; further data are available from the corresponding author upon reasonable request.

## Contribution Statement

AA was responsible for data collection, analysis and data interpretation. YA, AN and H.H.M contributed to the study conception and design. MKAK assisted with CT-Expo dose analysis for trunk CT examinations. FA contributed to data interpretation and drafted the manuscript. KM provided senior interpretive oversight, led manuscript development, integrated findings across sections and critical revision for scientific rigour.

## References

1. Brenner, D.J. and H.E. J., Computed tomography - An increasing source of radiation exposure. New England Journal of Medicine, 2007. 357(22): p. 2277–2284.

2. Mettler, F.A., et al., Effective doses in radiology and diagnostic nuclear medicine: A catalogue. Radiology, 2008. 248(1): p. 254–263.

3. United Nations Scientific Committee on the Effects of Atomic, R., Report of the United Nations Scientific Committee on the Effects of Atomic Radiation 2010, Fifty-seventh session, includes Scientific Report: summary of low-dose radiation effects on health. 2011.

4. Smith-Bindman, R., et al., Radiation Doses in Consecutive CT Examinations from Five University of California Medical Centres. Radiology, 2015. 277(1): p. 134–141.

5. Silva, A.C., et al., Innovations in CT Dose Reduction Strategy: Application of the Adaptive Statistical Iterative Reconstruction Algorithm. American Journal of Roentgenology, 2010. 194(1): p. 191–199.

6. Klink, T., et al., Reducing CT Radiation Dose with Iterative Reconstruction Algorithms: The Influence of Scan and Reconstruction Parameters on Image Quality and CTDIvol. European Journal of Radiology, 2014. 83(9): p. 1645–1654.

7. Kalra, M.K., A.D. Sodickson, and M.-S.W. W., CT Radiation: Key Concepts for Safe and Effective Use. RadioGraphics, 2015. 35(6): p. 1706–1721.

8. International Commission on Radiological, P., The 2007 Recommendations of the International Commission on Radiological Protection (Publication No. 2007, 103).

9. Wall, B. and S. P., The Historical Development of Reference Doses in Diagnostic Radiology. Radiation Protection Dosimetry, 1998. 80(1): p. 15–19.

10. Foley, S.J., M.F. McEntee, and R.L. A., Establishment of CT diagnostic reference levels in Ireland. The British Journal of Radiology, 2012. 85(1018): p. 1390–1397.

11. Malik, M.M.U.D., et al., An analysis of computed tomography diagnostic reference levels in India compared to other countries. Diagnostics, 2024. 14(15): p. 1585.

12. Leitz, W. and A. A., The impact of diagnostic reference levels on patient doses from X-ray examinations. 2008.

13. European, C., Diagnostic Reference Levels in Thirty-Six European Countries. 2014.

14. Brink, J.A. and M.D. L., U.S. National Diagnostic Reference Levels: Closing the Gap. Radiology, 2015. 277(1): p. 3–6.

15. International Commission of Radiation, P., Diagnostic Reference Levels in Medical Imaging (Publication No. 2017, 135).

16. McCollough, C.H., Diagnostic Reference Levels. 2010.

17. Wall, B.F., et al., Radiation risks from medical X-ray examinations as a function of patient age and sex (Report HPA-CRCE-028). 2011.

18. Protection, I.C.o.R., 1990 Recommendations of the International Commission on Radiological Protection (Superseded by ICRP Publication 103): Adopted by the Commission in November 1990: User’s Edition. 1992: International Commission on Radiological Protection.

19. Obed, R.I., G.I. Ogbole, and S.B. Majolagbe, Comparison of the ICRP 60 and ICRP 103 Recommendations on the Determination of the Effective Dose from Abdominopelvic Computed Tomography. International Journal of Medical Physics, Clinical Engineering and Radiation Oncology, 2015. 4(2): p. 172–176.

20. Christner, J.A., J.M. Kofler, and C.H. McCollough, Estimating effective dose for CT using dose–length product compared with using organ doses: consequences of adopting International Commission on Radiological Protection Publication 103 or dual-energy scanning. American Journal of Roentgenology, 2010. 194(4): p. 881–889.

21. Wardlaw, G., N. Martel, and H.C. Clinical Radiation Protection Bureau, Canadian Computed Tomography Survey National Diagnostic Reference Levels. 2016.

22. Qurashi, A.A., L.A. Rainford, and F.S. J., Establishment of diagnostic reference levels for CT trunk examinations in the western region of Saudi Arabia. Radiation Protection Dosimetry, 2014. 167(4): p. 569–575.

23. Shrimpton, P.C., et al., Doses from Computed Tomography (CT) Examinations in the UK - 2011 Review. 2014.

24. Bolowia, N., Establishment of computed tomography diagnostic reference levels in Tobruk. 2018.

25. Hayton, A., et al., Australian Diagnostic Reference Levels for Multi-Detector Computed Tomography. Australasian Physical & Engineering Sciences in Medicine, 2013. 36(1): p. 19–26.

26. Liang, C.R., et al., Establishment of an institutional diagnostic reference level for computed tomography with automated dose-tracking software. Journal of Medical Radiation Sciences, 2017. 64(2): p. 82–89.

27. American College of, R., ACR-AAPM-SPR PRACTICE PARAMETER FOR DIAGNOSTIC REFERENCE LEVELS AND ACHIEVABLE DOSES IN MEDICAL X-RAY IMAGING. 2018.

28. Yonekura, Y., Diagnostic reference levels based on latest surveys in Japan. Japan DRLs Report, 2015.

29. Abdalla, B., et al., The growing burden of cancer in the Gaza Strip. The Lancet Oncology, 2019. 20(8): p. 1054–1056.

30. Deevband, M.R., et al., Body-mass index-based effective dose determination in commonly performed computed tomography examinations in adults. Frontiers in Biomedical Technologies, 2022.

31. Niiniviita, H., et al., Dose monitoring in pediatric and young adult head and cervical spine CT studies at two emergency duty departments. Emergency Radiology, 2018. 25(2): p. 153–159.

32. Smith-Bindman, R., et al., International variation in radiation dose for computed tomography examinations: prospective cohort study. Bmj, 2019. 364: p. k4931.

33. Gnannt, R., et al., Automated tube potential selection for standard chest and abdominal CT in follow-up patients with testicular cancer: Comparison with fixed tube potential. 2012, European Radiology, 22(9), 1937–1945.

34. Beeres, M., et al., Chest-abdomen-pelvis CT for staging in cancer patients: Dose effectiveness and image quality using automated attenuation-based tube potential selection. Cancer Imaging, 2014. 14(1): p. 28.

35. Lira, D., et al., Tube potential and CT radiation dose optimisation. American Journal of Roentgenology, 2015. 204(1): p. W4–W10.

36. Mahesh, M., et al., Dose and pitch relationship for a particular Multislice CT scanner. American Journal of Roentgenology, 2001. 177(6): p. 1273–1275.

37. Kalra, M.K., et al., Comparison of Z-axis automatic tube current modulation technique with fixed tube current CT scanning of abdomen and pelvis. Radiology, 2004. 232(2): p. 347–353.

