## Supplementary materials for "Establishment of Local Diagnostic Reference Levels for Trunk Computed Tomography Examinations at Governmental Hospitals in the Gaza Strip: A Cross-Sectional Study"

**S1: Self-Administered Canadian CT Dose Survey Booklet:**

| Scanner Information | | | |
|---|---|---|---|
| CT Manufacture Company: | | Model: | |
| Instillation Date: | | Manufactured Date: | |
| There is a Periodic Maintenance? | Yes ☐<br>No ☐ | If Yes, How is it done? | - Yearly ☐<br>-Weakly ☐<br>-Monthly ☐<br>-Accidentally ☐ |
| Units Features Available: | Helical Scanning | Yes ☐ | No ☐ |
| | Max. Detector Configuration | | |
| | Dose Reduction Technique | Yes ☐ | No ☐ |
| Unit Type: | Single Slice CT (SSCT) ☐<br>Multi Slices CT (MSCT) ☐ | | |
| Other Usage: | Virtual Colonoscopy | Yes ☐ | No ☐ |
| | Interventional Procedures | Yes ☐ | No ☐ |
| | CT Angiography | Yes ☐ | No ☐ |
| Location of CT Unit: | ☐ Diagnostic Imaging Department | | |
| | ☐ Emergency Department | | |
| | ☐ Other (specify) | | |

### Individual Patient Survey

**Hospital Name:** ______ **Date:** ______

**Patient Number:** ______ **Examination:** Routine CT Trunk

- How many times he/she underwent for CT trunk examination? ______

-When was the last time this patient underwent this examination? ______

| | | | |
|---|---|---|---|
| **Patient Information** | **Age(y)** | | |
| | **Gender(M/F)** | | |
| **Anthropometric Data** | **Weight(kg)** | | |
| | **Height(cm)** | | |
| | **Phase 1** | **Phase 2** | **Phase 3** |
| **Indicate actual start and end positions with lines on each image** → | 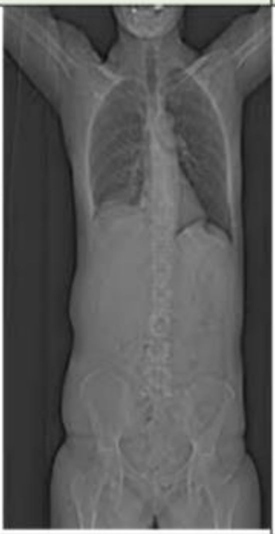 | 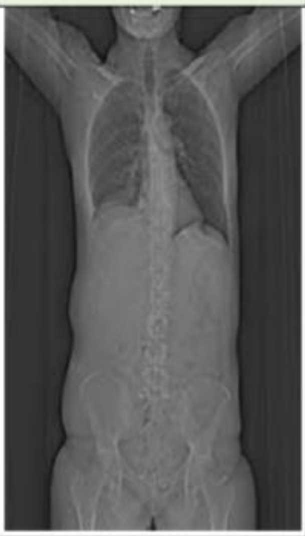 | 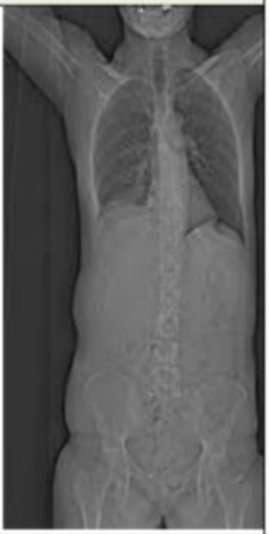 |
| **Describe anatomical range scanned Scanned range (cm)** | | | |
| **Z-(start)** | | | |
| **Z+(end)** | | | |
| **Scan Range (cm)** | | | |
| **Sureview AP./ LAT.** | | | |
| **I.V Contrast? Indicate Phase Name?** | Yes ☐ No ☐ | Yes ☐ No ☐ | Yes ☐ No ☐ |
| **Detector Configuration** | | | |
| **Tube Voltage (Kv)** | | | |
| **Tube Rotation Time (s)** | | | |

| | | | | |
|---|---|---|---|---|
| Tube Current (mA) | | | | |
| Displayed mA . s<br>-mAs ☐<br>-mAs/Slice ☐<br>-effective mAs ☐ | | Average mAs | Average mAs | Average mAs |
| Auto-Dose Reduction Used ?<br>Name? | | Yes ☐ No ☐ | Yes ☐ No ☐ | Yes ☐ No ☐ |
| Axial Scanning | Helical Scanning | Axial ☐ Helical ☐ | Axial ☐ Helical ☐ | Axial ☐ Helical ☐ |
| No. of axial slices | Scan Length (cm) | | | |
| Table Increment (mm) | Pitch | | | |
| Over scan or partial scan angle(+ or -) | Table Speed/Travel (mm per rotation) | | | |
| Console CTDIvol | | | | |
| Console DLP-Sequence | | | | |
| Console DLP-Exam | | | | |
| hrec/ Reconstructed Slice Thickness (mm) | | | | |
| Number of Slices | | | | |
| hcol | | | | |
| N*hcol | | | | |
| Gantry Tilt | | | | |
| Number of Series | | | | |
| Shielding Type Used? | | Lead ☐ | | Bismuth ☐ |
